# A blood-based signature of cytoskeletal and extracellular remodeling for risk stratification of intraductal papillary mucinous neoplasms

**DOI:** 10.64898/2026.08.09.26360008

**Authors:** LaNisha Patterson, Riccardo Ballarò, Yihui Chen, Ana Vilchis Celis, Mingxin Zuo, Benson Chellakkan Selvanesan, Alejandra Flores Villanueva, Ehsan Irajizad, Eugene Koay, Michael P. Kim, Cynthia Reinhart-King, Thoa Tran, Anirban Maitra, Jianjun Zhang, C Max Schmidt, Samir Hanash, Johannes F. Fahrmann

**Author notes:** Corresponding Author: Johannes F. Fahrmann, PhD, The University of Texas MD Anderson Cancer Center 6767 Bertner Street, Houston, TX 77030, USA.

## Abstract

**Background:** Intraductal papillary mucinous neoplasms (IPMNs) are recognized as precursor lesions to pancreatic ductal adenocarcinoma (PDAC). However, the molecular programs underlying progression from low-grade dysplasia to advanced disease remain incompletely characterized. Herein, we performed an integrated plasma and tissue–proteomic analyses coupled with spatial and single-cell transcriptomics to identify biologically coherent remodeling programs reflected in circulation that distinguish IPMN by dysplasia grade and invasive disease.

**Methods:** Using the O-link proximity extension assay platform, a panel of 1,104 proteins were quantified in plasma samples collected from patients with low-grade (LG) IPMN (n=30), high-grade (HG) IPMN with or without associated PDAC (IPMN/PDAC; n=40) and PDAC without IPMN (n=8). Predictive performance of individual biomarkers were assessed; likelihood ratio testing was performed to identify protein biomarkers that were complementarity with CA19-9 for risk of malignancy of IPMN. Findings were intersected with available spatial (N= 13) and single-cell (N= 6) transcriptomic datasets of IPMN tissues as well as mass spectrometry-based proteomic profiles of an independent set of resected human IPMN tissues (N= 9).

**Results:** A total of 28, 43, and 35 circulating proteins were found to be differential in HG, IPMN/PDAC, and HG + IPMN/PDAC cases compared to LG IPMN. Among differential proteins were known PDAC-associated markers CEACAM5, CTRC, and REG3A as well as several biomarkers reflecting cytoskeletal and extracellular matrix remodeling and inflammatory processes. Focusing on cytoskeletal and ECM-related proteins and using likelihood ratio testing, an “OR” rule considering CA19-9, BGN, and ITGB1BP1 achieved overall sensitivity of 48.7% for HG + IPMN/PDAC, including 38.1% sensitivity for HG IPMN, at an overall specificity of 90%, which was improved compared to that of CA19-9 alone (overall sensitivity of 28.2%; McNemar Exact test 1-sided p-value: 0.011). Integrated proteomic and spatial transcriptomic datasets of IPMN tissues revealed coordinated alterations cytoskeletal and ECM remodeling and elevated matrix stiffness as prominent features associated with IPMN/PDAC, which paralleled concordant increases in BGN and ITGB1BP1. Cell-type of origin analyses based on spatial and single-cell data further revealed fibroblasts and myeloid cells as primary contributors to expression levels of BGN whereas ITGB1BP1 was primarily expressed in neoplastic epithelium.

**Conclusion:** Advanced IPMN dysplasia and invasive disease are characterized by coordinated tissue remodeling programs that are systemically reflected in circulating proteomic profiles. Blood-based biomarkers identified through our study, such as BGN and ITB1BP1, have potential to improve upon CA19-9 for risk stratification of IPMN to better guide clinical management.

## Introduction

Pancreatic cysts occur in up to 13% of patients undergoing abdominal imaging (CT scan or MRI) for reasons unrelated to pancreatic symptoms[1]. Although pancreatic cysts are frequent with a prevalence of upwards of 24% [1–3], most of these cysts will not ultimately progress to pancreatic cancer. The most common cystic neoplasm that is an established precursor to pancreatic ductal adenocarcinoma (PDAC) is intraductal papillary mucinous neoplasm (IPMN). IPMN is a mucin-secreting neoplasm arising in either the main pancreatic duct or one of the branch ducts and comprises nearly half of resected lesions that are initially diagnosed as asymptomatic pancreatic cysts [1, 4–6]. IPMNs are lined by either low-grade (LG) or high-grade (HG) epithelial dysplasia, and, for a subset of cases, histological progression culminates in PDAC where the probability of survival is drastically reduced [1, 4–6].

International consensus guidelines recommend either resection of IPMN with high risk of malignancy or surveillance of IPMN without surgical indications [7]. While current guidelines achieve satisfactory sensitivity (>90%), they are hindered by limited specificity (25-30%) for predicting malignant IPMN compared with surgical pathology [8, 9]. The low specificity leads to potential of overdiagnosis, which is associated with high morbidity (20-40%) and possibility of mortality (1-5%) in patients with benign (low- or moderate-grade) IPMN [10, 11].

It is well recognized that IPMN progression from low-grade dysplasia to invasive disease is not solely driven by epithelial transformation but is potentiated by remodeling of the tumor microenvironment. These changes include alterations in stromal composition, immune cell infiltration, and extracellular matrix (ECM) organization, together with shifts in secretory signaling networks that promote malignant progression. Importantly, coordinated microenvironmental reprogramming is reflected in circulating protein profiles, providing opportunity for development of blood-based biomarkers for predicting risk of malignancy of IPMN and better guiding clinical management.

In the current study, we applied Olink proximity extension assay technology to establish proteomic signatures associated with dysplasia grade and invasive disease in IPMN. Specifically, we profiled an “off the shelf” panel of 1,104 proteins in plasma from a total of 78 patients with LG IPMN (n = 30), HG IPMN with or without associated PDAC (n = 40), or PDAC (n = 8). Findings were further integrated with mass spectrometry-based proteomic profiles of an independent set of resected human IPMN tissues (N= 9) as well as available spatial and single-cell transcriptomics datasets to interrogate spatiomolecular distributions and infer cell-type of origin for candidate biomarkers. The complementarity of protein biomarkers with CA19-9 for improved risk stratification was also evaluated.

## Methods

### Plasma Samples

Plasma specimens were obtained from a total of 78 patients undergoing clinical evaluation or surgical resection for pancreatic cystic disease at Indiana University School of Medicine under institutional review board–approved protocols [12]. The specimen set comprised of 30 plasma samples form patients with LG IPMN, 21 patients with HG IPMN, and 19 patients with HG and an associated PDAC (IPMN/PDAC). An additional 8 plasma samples were included from PDAC patients without an associated IPMN **(Table 1)**. Eight additional randomized duplicate samples were included to assess technical reproducibility. Cases and controls were frequency-matched based on age, sex, diabetes status, smoking history, and alcohol use (**Table 1**).

**Table 1.**
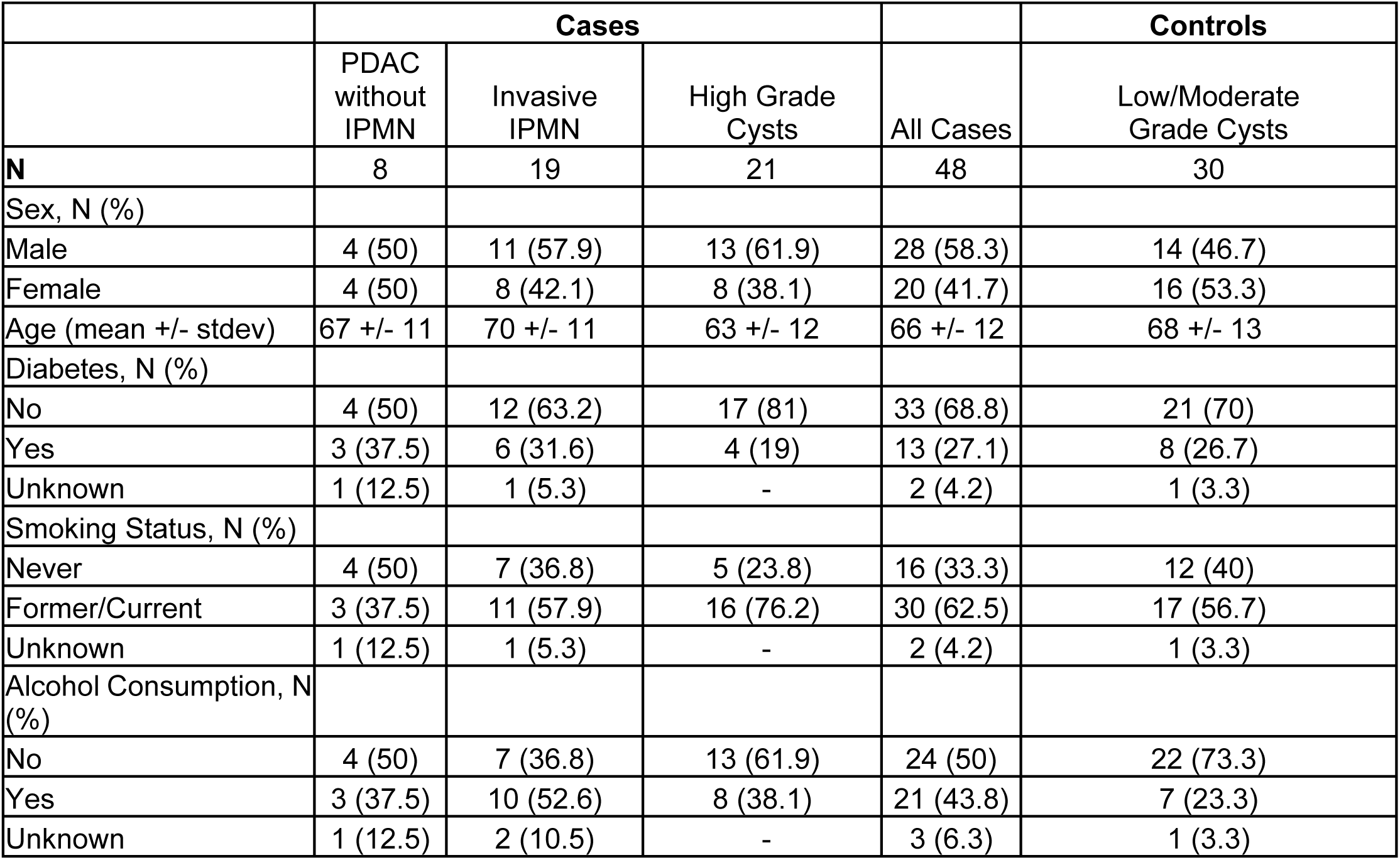
Patient and tumor characteristics.

### IPMN Tissues

Flash-frozen human resected IPMN tissues were derived from the MD Anderson Pancreas Tissue & Blood Biobank under IRB approved protocol LAB00–396 in accordance with the Declaration of Helsinki. The specimen set consisted of 3 IPMN tissues with LG dysplasia, 3 with HG dysplasia, and 3 with HG dysplasia with a concurrent PDAC (IPMN/PDAC), all histologically validated in resection samples [12, 13].

### O-link plasma proteomic profiling

The O-link proximity extension assay platform was used to quantify 1,104 proteins circulating proteins in plasma of patients with IPMN (**Supplemental Table S1**). Approximately 25–30 µL of plasma was used per sample in accordance with manufacturer recommendations. Normalized protein expression (NPX) values were obtained using the standard O-link normalization workflow [14].

### Mass Spectrometry Analysis of IPMN Tumor Tissue

Quantitative proteomic analysis of low-grade IPMN, high-grade IPMN, and IPMN-associated PDAC tumor tissues was performed as previously described by Ballarò et al. [13]. Briefly, 0.5ug of peptides were loaded onto a pre-conditioned EvoTip and peptides separated using an Evosep One chromatography system coupled to a Bruker timsTOF HT mass spectrometer operating in diaPASEF acquisition mode. Mass spectrometry data were processed using Spectronaut against the UniProt SwissProt human reference proteome with peptide and protein identifications filtered at a 1% false discovery rate (FDR). Relative protein abundance across samples was determined using label-free quantification approaches [13].

### Spatial and single-cell transcriptomic datasets of IPMN tissues

Spatial transcriptomic (ST) data (Visium 10X platform) was derived from our recent publication[12, 15]. Spots in the ST dataset were annotated into three regions: epilesional (Epi), juxtalesional (Juxta), and perilesional (Peri). Epilesional was defined as areas covering the epithelial lining, juxtalesional as adjacent microenvironment to the lesion, and perilesional referring as an additional layer of stroma outside of the epithelium [12, 15]. Single-cell data integration method to match and compare (scMC) pipeline was used for initial processing and clustering of the human IPMN ST dataset. To analyze spatially-resolved RNA-seq data, we used the Seurat package (version 5.5.0) implemented in R statistical software (version 4.5.2) (https://www.r-project.org/) [12]. A compositive matrix stiffness score was derived through summation of transcript expression levels of core genes associated with matrix stiffness (**Supplemental Table S2**). Cell type-specific expression profiles were derived using robust cell-type deconvolution (RCTD) algorithms as described previously [16, 17].

### Ingenuity Pathway Analysis

Pathway enrichment analysis was performed using Ingenuity Pathway Analysis (IPA; Qiagen). Statistical significance of enriched pathways was determined using a right-tailed Fisher’s exact test.

### Statistical analysis

For human plasma analyses, Area under the receiver operating characteristic curve (AUCs) were determined for individual protein biomarkers for differentiating advanced neoplasia (HG IPMN, IPMN/PDAC, and PDAC) from LG IPMN using the pROC package in R statistical software (version 4.5.2) (https://www.r-project.org/) [12].

The 95% confidence intervals for each biomarker were determined using a bootstrap procedure wherein data was re-sampled with replacement separately for the controls and the diseased 1,000 bootstrap samples. Statistical significance was assessed using Wilcoxon rank-sum tests and 2-sided p-values reported. To account for multiple testing, p-values were adjusted using the Benjamini–Hochberg method. Go Biological Processes were determined using the ‘ClusterProfiler’, ‘org.Hs.eg.db’, and ‘enrichplot packages’ in R statistical software (version 4.5.2). Likelihood ratio testing of individual protein biomarkers with CA19-9 was performed R statistical software (version 4.5.2). McNemar exact tests were used to compare two binomial proportions of IPMN patients with two different biomarker scores as previously described [17]. For proteomic analysis of IPMN tissues, proteins below limit-of-detection were imputed with +1 and values log2 normalized prior to statistical analysis and statistical significance determined using 2-sided Student T-tests. Figures were generated using R statistical software (version 4.5.2) and GraphPad Prism (version 10.6.1).

## Results

### Plasma proteomic signatures of cytoskeletal and extracellular matrix remodeling are associated with dysplasia grade and invasive disease in IPMN

To identify differential circulating proteins associated with higher dysplasia grade and invasive disease in IPMN, four comparative analyses were performed: HG IPMN vs LG IPMN, IPMN/PDAC vs LG IPMN, HG + IPMN/PDAC vs LG IPMN, and PDAC without an associated IPMN vs LG IPMN. Statistical analyses revealed 28, 43, 35, and 185 proteins to be differential (nominal Wilcoxon rank sum test 2-sided p-value < 0.05) across the respective comparative groups (**Figure 1A; Supplemental Table S1**). Among proteins associated with HG IPMN with or without an associated PDAC (IPMN/PDAC) were CEACAM5, CTRC, and REG3A, which are known biomarkers associated with PDAC (**Supplemental Table S1**).

**Figure 1.**
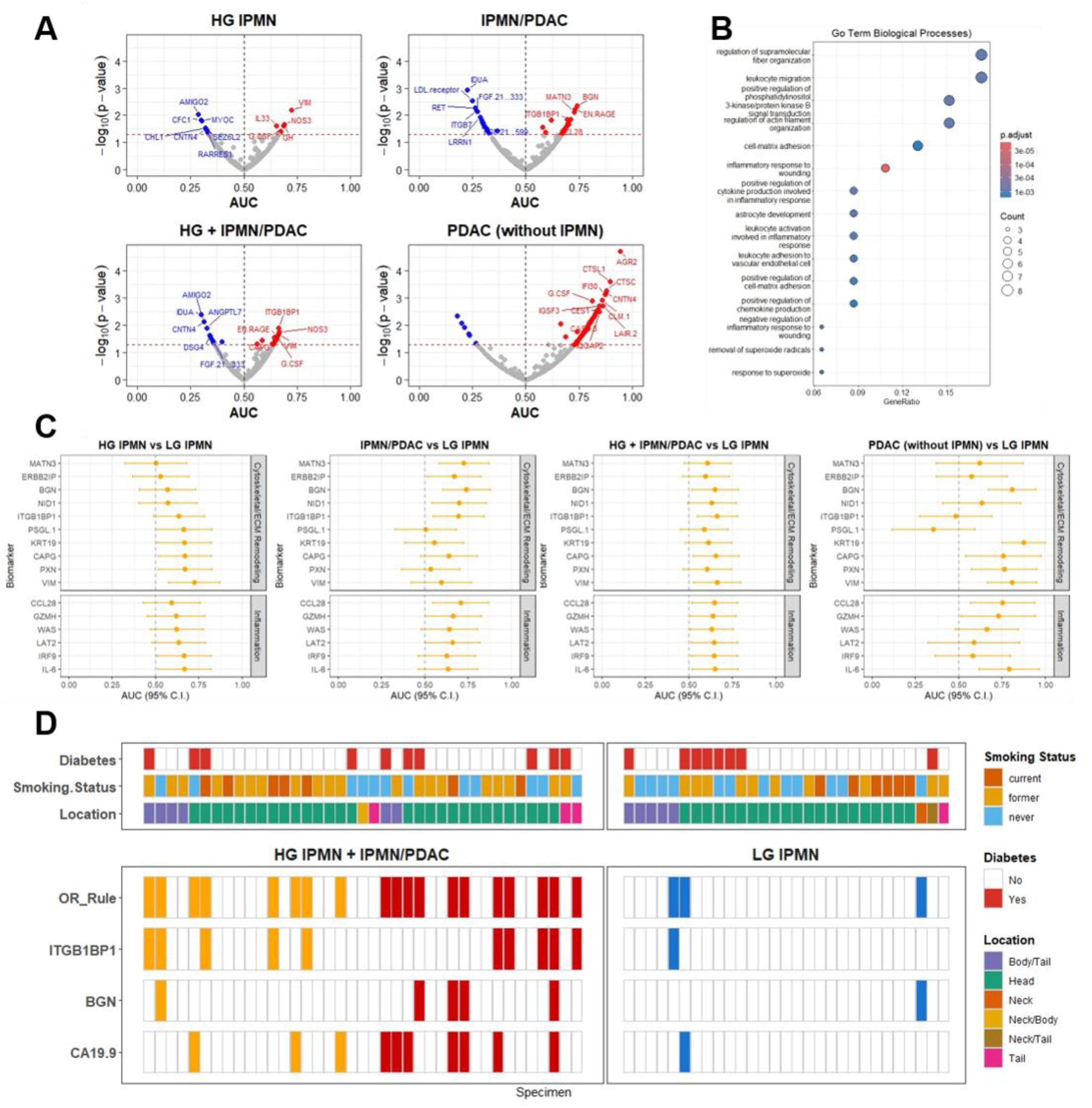
Circulating protein signatures of cytoskeletal and extracellular remodeling and inflammatory processes are association with dysplasia grade and invasive disease progression of IPMN. **A)** Volcano plots demonstrate differential circulating proteins when considering HG IPMN, IPMN/PDAC, HG + IPMN/PDAC, or PDAC without an associated IPMN compared to LG IPMN. **B)** Go Biological Process enrichment analysis based on differentially elevated (AUC >0.5; p-value <0.05) proteins in HG IPMN with or without an PDAC compared to LG IPMN. **C)** Dot plots showing Area under the Receiver Operating Characteristic Curve (AUC) and 95% Confidence Intervals for circulating proteins that reflect cytoskeletal and extracellular remodeling (ECM) or inflammation when considering HG IPMN vs LG IPMN, IPMN/PDAC vs LG IPMN, HG IPMN + IPMN/PDAC vs LG IPMN, and PDAC without an associated IPMN vs LG IPMN. **D)** ‘OR’-rule approach for individual protein biomarkers for rule-in applications based on individual marker 95% specificity thresholds.

Go Biological Process enrichment analysis of protein biomarkers elevated (AUC > 0.5; p-value<0.05) in cases (HG IPMN or IPMN/PDAC) demonstrated enrichment of cytoskeletal and extracellular matrix (ECM) remodeling as well as inflammatory processes (**Figure 1B-C; Supplemental Table S1**). Several of these markers, including BGN, CAPG, KRT19, MATN3, NID1, PXN, and VIM, were also found to be elevated (AUC > 0.60) in plasma from PDAC cases without an associated IPMN (**Supplemental Table S1**), indicating a biological network more broadly associated with PDAC.

In the IPMN cohort, CA19-9 had an AUC of 0.76 (95% CI: 0.61-0.91) for distinguishing IPMN/PDAC from LG IPMN with limited performance for HG IPMN (**Supplemental Figure S1**). Likelihood ratio testing identified several protein biomarkers, including CEACAM5 as well as cytoskeletal and ECM-related proteins KRT19, BGN and ITGB1BP1, to be complementary (1-sided p-value <0.05) to CA19-9 for improved predictive performance (**Supplemental Table S2**). Focusing on cytoskeletal and ECM-related proteins, an ‘OR’ rule considering CA19- 9, BGN, and ITGB1BP1 at individual biomarker thresholds corresponding to 95% specificity achieved an overall sensitivity of 48.7% with an overall specificity of 90% for HG + IPMN/PDAC, which was improved compared to a sensitivity of 28.2% for CA19-9 alone at equivalent 90% specificity (McNemar exact test 1-sided p-value: 0.011) (**Figure 1D**). Of relevance, the ‘OR’-rule had a sensitivity of 38.1% for HG IPMN, compared to sensitivity of 9.5% for CA19-9 alone (McNemar exact test 1-sided p-value: 0.031) (**Figure 1D**). Collectively, these data support protein signatures indicative of cytoskeletal and ECM re-modeling as potential biomarkers for predicting risk of malignancy of IPMN.

### Integrated proteomics and spatial transcriptomics of IPMN tissues reveal cytoskeletal and ECM remodeling and elevated matrix stiffness as a prominent feature associated with progression of IPMN

We considered that the observed increases in circulating cytoskeletal/ECM-related protein biomarkers (e.g. BGN and ITGB1BP1) (**Figure 1**) may reflect alterations in pancreas tissue architecture and matrix stiffness that associated with disease progression [18, 19]. To evaluate this hypothesis, we first compared proteomic profiles generated on an independent set of human IPMN tissues (3 LG, 3 HG, and 3 IPMN/PDAC). Consistent, analyses revealed several key proteins involved in cytoskeleton and ECM remodeling, including BGN and ITGB1BP1, and matrix stiffness to be variably elevated (median fold change > 1.25) in HG and IPMN/PDAC compared to LG IPMN (**Supplemental Table S3; Figure 2A**).

**Figure 2.**
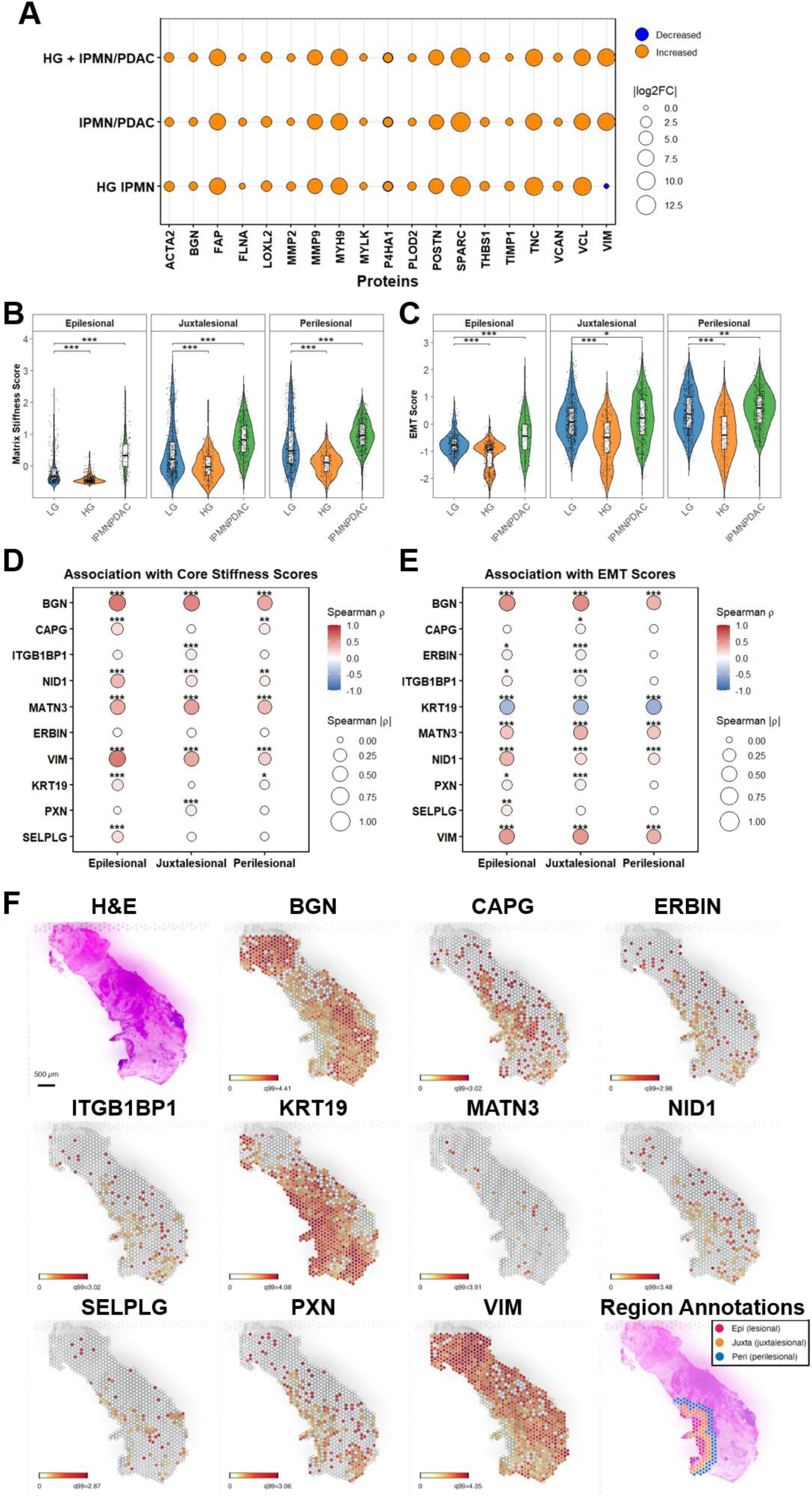
Association between gene signatures related to cytoskeletal and ECM remodeling and matrix stiffness and dysplasia grade and invasive disease progression of IPMN. **A)** Bubble plot illustrates median fold-change in cytoskeletal/ECM and matrix stiffness related proteins in HG IPMN and IPMN/PDAC tissues compared to LG IPMN tissues (N= 3 per group). **B-C)** Composite matrix stiffness (**B**) and EMT scores (**C**) in LG IPMN, HG IPMN, and IPMN/PDAC tissues based on spatial transcriptomic data. **D-E)** Bubble plot depicting spearman rho correlation coefficients between mRNA expression levels of respective genes and compositive matrix stiffness (**D**) and EMT (**E**) scores. 2-sided p-values *<0.05; **<0.01; ***<0.001. **F)** Spatial transcriptomic images demonstrate spatial distribution of mRNA expression for respective genes in epilesional, juxtalesional, and perilesional regions.

Next, we leveraged a previously reported Visium 10X spatial transcriptomic dataset generated from 13 resected human IPMN tissues (N = 7 LG IPMN, 3 HG IPMN, and 3 IPMN/PDAC) to assess spatiomolecular patterns of transcriptional programs related to tissue remodeling and matrix stiffness across IPMN dysplasia grades and invasive disease. Gene Set Enrichment Analysis (GSEA) of differentially expressed genes (DEGs; FDR-adjusted p-value < 0.05) revealed significant enrichment of hallmark pathways associated with cytoskeletal and ECM remodeling, including epithelial-to-mesenchymal transition (EMT), TNFα signaling via NF-κB, and apical junction pathways, in HG IPMN and IPMN/PDAC tissues compared with LG IPMN tissues (Supplemental Figure S2A). Moreover, composite EMT and matrix stiffness scores were markedly higher in IPMN/PDAC compared with LG IPMN, whereas HG IPMN exhibited lower composite EMT and matrix stiffness scores (Figure 2B-C). These data indicate dynamic changes in tissue-remodeling and matrix stiffness-associated transcriptional programs across IPMN dysplasia grade and invasive disease, with the most pronounced alterations observed in IPMN-associated PDAC.

Spearman correlation analyses comparing EMT and matrix stiffness scores with mRNA expression levels of BGN and ITG1BP1 are shown in **Figures 2D-E** and in **Supplemental Figures S2B, S3 and S4**. For broader interrogation, we additionally considered other cytoskeletal/ECM-related protein biomarkers that were associated with HG IPMN and IPMN/PDAC (**Figure 1**). Analyses revealed mRNA expression of BGN in addition to NID1, MATN3, PSGL-1/SELPLG, and VIM to be positively correlated with EMT and matrix stiffness scores, particularly in epilesional regions. Gene expression of CAPG and KRT18 positively correlated with matrix stiffness scores and inversely associated with EMT scores; whereas ITGB1BP1 and PXN mRNA expression were inversely associated with both EMT and matrix stiffness scores. Although ITGB1BP1 demonstrated negative correlations with EMT and matrix stiffness scores, mRNA expression and protein levels of ITGB1BP1 were consistently observed to be higher in HG IPMN and IPMN/PDAC compared to LG IPMN tissues (**Supplemental Figure 2B; Supplemental Table S3**). Functionally, ITGB1BP1, also known as Integrin Cytoplasmic Domain-Associated Protein 1 (ICAP1), serves as a mechanotransducer that binds B1 integrin and aids in ECM remodeling and matrix stiffness, contributing to metastatic potential of tumor cells [20, 21]. Similarly, PXN is also involved in mechano-transducing machinery, serving as a substrate for activating β3 integrin-talin1- kindlin [22].

### Cell-types of origin for candidate protein biomarkers inferred through spatial and single-cell transcriptomic analyses of IPMN tissues

Stratification by epiliesional, juxtalesional, and perilesional regions demonstrated mRNA expression of BGN, and VIM to be predominately expressed in juxtalesional and perilesional regions whereas CAPG, ERBB2IP/ERBIN, ITGB1BP1, and PXN transcripts were enriched in epilesional areas (**Figure 2F; Supplemental Figure S2C**). KRT19, MATN3, NID1, and PSGL-1/SELPLG were broadly expressed in all three regions (**Figure 2F; Supplemental Figure S2C**). To interrogate cell-type(s) of origin for above mentioned candidates, we leveraged available single-cell transcriptomic datasets generated on an independent set of IPMN tissues (N= 2 LG IPMN; 2 HG IPMN; 2 IPMN/PDAC). As shown in representative t-SNE plots, a total of 8 cell-types were inferred (**Figure 3A; Supplemental Figure S5**). Consistent with prior reports, an alluvial plot depicting distribution of cell-type populations demonstrated a shift towards increased intratumor presence of fibroblasts, macrophages, dendritic cells, and other myeloid cells in HG IPMN and IPMN/PDAC compared to LG IPMN [23–25] (**Figure 3B**). Expression levels of BGN, MATN3, and NID1 were found to be prominent in fibroblasts whereas KRT19 was primarily expressed in epithelial cells (**Figure 3C**). CAPG, ITGB1BP1, PSGL-1/SELPLG, and VIM were highly expressed in epithelial and immune cell subtypes whereas PXN was broadly expressed (**Figure 3C**). ERBB2IP/ERBIN was not identified in the single-cell dataset. Notably, expression profiles of respective genes increased in respective cell-type compartments with progression to HG IPMN and IPMN/PDAC (**Figure 3D**). Collectively, these results demonstrate coordinated remodeling across the neoplastic epithelium and surrounding stromal and immune microenvironment.

**Figure 3.**
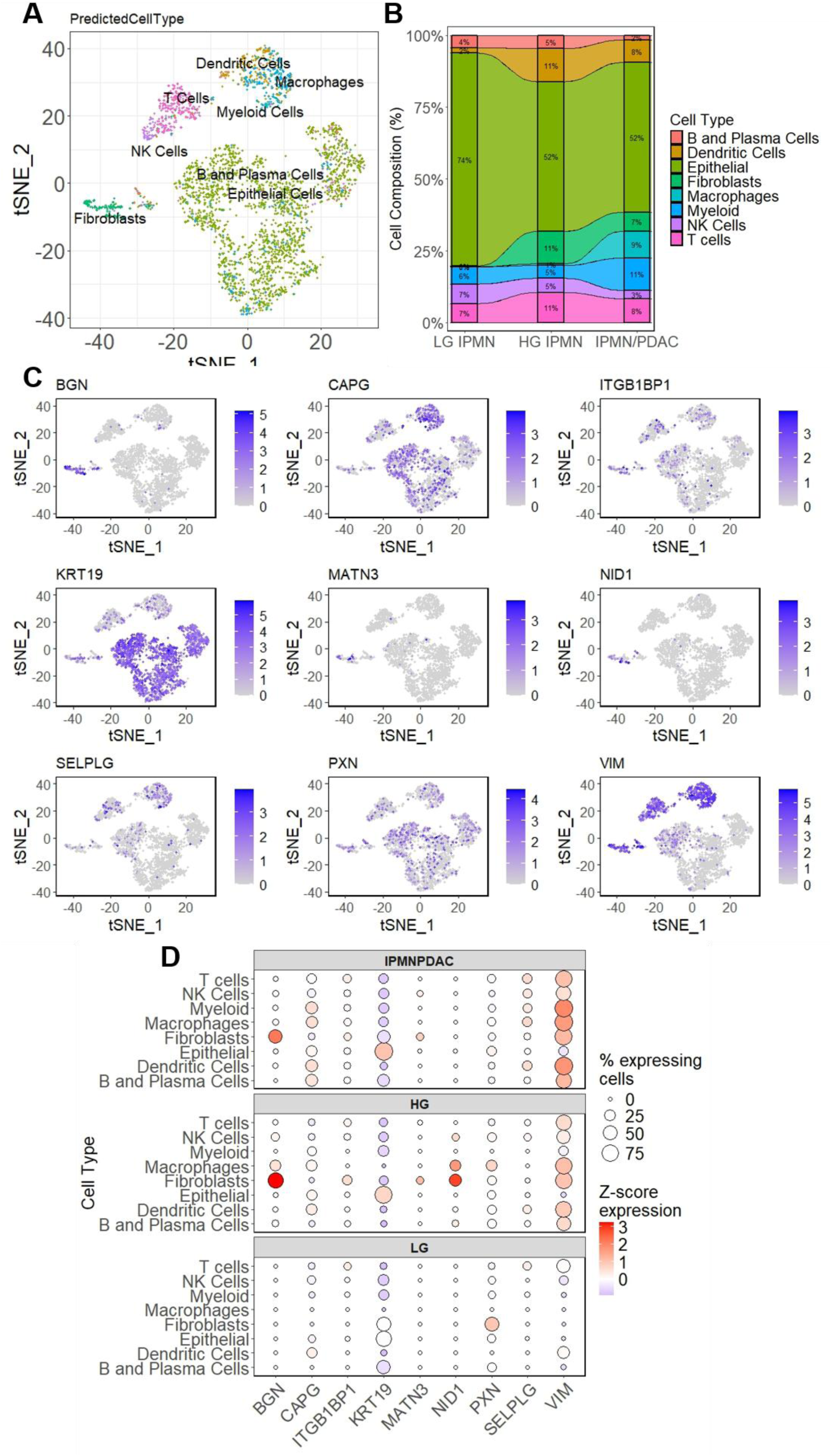
Cell-types of origin for candidate protein biomarkers inferred through single-cell transcriptomic analyses of IPMN tissues. **A)** Representative t-SNE plot illustrating 8 cell types inferred in the single-cell transcriptomic dataset of IPMN tissues (N= 2 LG IPMN, 2 HG IPMN, and 2 IPMN/PDAC). **B)** Alluvial plot illustrating changes in cell populations. **C)** representative t-SNE plots for BGN, CAPG, ITGB1BP1, KRT19, MATN3, NID1, PSGL-1/SELPLG, PXN, and VIM. ERBB2IP/ERBIN was not identified in the single-cell dataset. D) Bubble plots depicting expression levels of BGN, CAPG, ITGB1BP1, KRT19, MATN3, NID1, PSGL- 1/SELPLG, PXN, and VIM across different cell types when stratifying samples into LG IPMN, HG IPMN, and IPMN/PDAC.

## Discussion

Accurate identification of intraductal papillary mucinous neoplasms (IPMNs) at harboring high-grade dysplasia or invasive carcinoma remains a major clinical challenge. Although current consensus guidelines achieve high sensitivity for identifying high-grade dysplasia or invasive carcinoma, their limited specificity contributes to the surgical overtreatment of patients with low-grade or nonmalignant lesions, exposing them to the substantial morbidity associated with pancreatic resection. Moreover, CA19-9, the most used circulating biomarker in this setting, is more strongly associated with invasive carcinoma than with high-grade dysplasia and therefore has limited sensitivity for detecting advanced neoplasia before invasion occurs [26–28]. In the current study, we applied plasma proteomics with tissue proteomics, spatial transcriptomics, and single-cell transcriptomics to identify biologically informed circulating biomarkers for distinguishing high-grade IPMN with or without an associated PDAC from low-grade IPMN. These analyses led to the identification of a blood-based signature associated with higher dysplasia grade and invasive disease in IPMN and reflecting coordinated cytoskeletal, extracellular matrix remodeling, and inflammatory programs. Moreover, our study identified candidate targets, such as BGN and ITG1BP1, that are complementary to CA19-9 for improved risk stratification of IPMN.

Among circulating proteins associated with high-grade dysplasia and IPMN-associated PDAC were previously established pancreatic cancer–associated markers such as CEACAM5 [29], CTRC [30], KRT19 [31], and REG3A [32], as well as proteins involved in cytoskeletal organization, extracellular matrix remodeling, and inflammatory signaling. To this end, BGN, CAPG, ERBB2IP/ERBIN, ITGB1BP1, MATN3, NID1, PSGL- 1/SELPLG, PXN, and VIM emerged as prominent components of a tissue-remodeling signature associated with advanced neoplasia. The biological relevance of this circulating signature was further supported by integrated tissue proteomic and spatial transcriptomic analyses. HG IPMN and IPMN-associated PDAC lesions exhibited coordinated increases in proteins and transcriptional programs associated with cytoskeletal organization, extracellular matrix remodeling, cellular contractility, and matrix stiffness. These alterations were accompanied by enrichment of epithelial-to-mesenchymal transition, TNFα/NF-κB signaling, and apical-junction pathways, with the most pronounced changes observed in IPMN-associated PDAC. Together, these findings suggest that the plasma signature reflects coordinated changes in tissue architecture and mechanobiology, including cytoskeletal reorganization, extracellular matrix remodeling, and altered matrix stiffness, that characterize higher-grade and invasive IPMN rather than isolated alterations in individual proteins.

Of the remodeling-associated candidates, BGN has been directly linked to extracellular matrix organization and TGF-β–regulated stromal signaling. Specifically, BGN is a small leucine-rich proteoglycan that participates in collagen fibrillogenesis, matrix assembly, growth-factor sequestration, and cell–matrix communication [31–33]. In pancreatic cancer models, BGN has been linked to TGF-β–SMAD4 and ERK signaling, EMT-associated phenotypes, and cellular motility [33]. In the present study, the concordant elevation of BGN in plasma and tissue, together with its positive association with EMT and matrix-stiffness scores, supports its potential utility as a circulating marker of ECM and microenvironmental remodeling associated with higher dysplasia grade and invasive disease. This biology is distinct from the epithelial glycan phenotype represented by CA19-9 and may provide a plausible explanation for the observed complementarity of BGN with CA19-9 for improved risk stratification of IPMN. In contrast, ITGB1BP1 may reflect altered β1-integrin–dependent cell–matrix signaling. ITGB1BP1 directly binds the cytoplasmic domain of β1 integrin and regulates integrin activation, cell adhesion, and cytoskeletal organization [34]. β1-integrin signaling is well established in pancreatic cancer progression, where it has been associated with tumor–matrix interactions, invasion, metastasis, recurrence, and adverse clinical outcomes [35]. Although the specific contribution of ITGB1BP1 to pancreatic neoplasia remains less well defined, its reported dysregulation in invasive PDAC and its consistent elevation in advanced IPMN lesions support its potential relevance to altered integrin-associated signaling in higher grade and invasive IPMN [36].

Spatial and single-cell transcriptomic analyses further revealed that the candidate biomarkers, including BGN and ITGB1BP1, reflect contributions from distinct epithelial and microenvironmental compartments. The increased representation of fibroblasts, macrophages, dendritic cells, and other myeloid populations in HG IPMN and IPMN-associated PDAC was consistent with prior single-cell studies demonstrating progressive remodeling of the stromal and immune microenvironment during IPMN progression [23, 37]. BGN, NID1, and VIM were preferentially expressed in juxtalesional and perilesional regions and associated with fibroblast and myeloid cell signatures. This distribution is consistent with the established role of cancer-associated fibroblasts in producing and reorganizing the extracellular matrix and in communicating with myeloid and other immune populations through matrix-dependent and paracrine signaling. The functional and spatial heterogeneity of cancer-associated fibroblasts can generate distinct stromal niches characterized by differences in matrix deposition, tissue mechanics, inflammatory signaling, and immune regulation [37]. In particular, the stromal localization of BGN is consistent with its established function as an extracellular matrix proteoglycan and its reported overexpression and TGF-β–dependent regulation in the desmoplastic microenvironment of pancreatic cancer [33, 38–40]. NID1, a structural component of basement membranes, has also been implicated in extracellular-matrix–dependent tumor growth and metastatic progression [41]. VIM is broadly expressed in mesenchymal and stromal populations, while its acquisition by pancreatic tumor cells is associated with epithelial-to-mesenchymal transition and more aggressive clinical behavior [42, 43]. Their perilesional localization may therefore reflect structural and phenotypic remodeling at the interface between the neoplastic lesion and surrounding tissue. Together, the coordinated expression of basement membrane–associated proteins such as NID1, mesenchymal markers such as VIM, epithelial cytoskeletal proteins including KRT19, and regulators of focal adhesion and cell–matrix interactions supports a tissue-remodeling program consistent with an invasion-permissive microenvironment. By contrast, CAPG and ITGB1BP1 were enriched within epilesional regions and showed stronger associations with epithelial signatures, suggesting a closer relationship with cytoskeletal remodeling and cell–matrix interactions within the neoplastic compartment. CAPG is an actin-capping protein that regulates cytoskeletal dynamics and has recently been shown to promote pancreatic ductal adenocarcinoma cell proliferation and migration, with elevated expression associated with adverse clinical outcomes [44]. ITGB1BP1 directly binds the cytoplasmic domain of β1 integrin and regulates integrin activation, focal-adhesion organization, cell adhesion, and migration [45, 46]. Analysis of an independent single-cell IPMN dataset further refined these spatial findings by demonstrating that the candidate proteins were not restricted to a single cellular population. BGN, NID1, VIM, and ITGB1BP1 were detected predominantly in fibroblasts, dendritic cells, and other myeloid populations, whereas CAPG was expressed in epithelial cells as well as stromal and immune compartments. Collectively, these observations suggest that the circulating signature does not arise from a single cellular source but instead captures coordinated epithelial, stromal, and immune remodeling associated with higher dysplasia grade and invasive disease.

Several limitations should be considered. The plasma, tissue proteomic, spatial, and single-cell analyses were based on limited datasets and previously collected specimen cohorts. The biomarker thresholds and three-marker classification approach were developed and evaluated within the same discovery cohort and therefore require validation in larger, independent, prospectively collected populations. Because samples were obtained at clinical evaluation or surgical resection, the current study cannot determine whether these biomarkers predict future progression during surveillance. Although integration across orthogonal platforms supports the biological relevance of the circulating candidates, their specific contributions to IPMN progression and the mechanisms underlying their release into circulation remain to be defined. Future studies should also determine whether these markers provide incremental value beyond established clinical, radiographic, and pathologic risk factors.

In summary, higher IPMN dysplasia grade and invasive disease are associated with coordinated cytoskeletal and extracellular matrix remodeling that is manifest in circulation. Blood-based biomarkers, such as BGN and ITGB1BP1, reflecting these biological processes may complement CA19-9 for improved risk stratification of IPMN to better guide clinical management.

## Data Availability

All data produced in the present study are available upon reasonable request to the authors

## Acknowledgements

A.M. is supported by NCI P50CA221707, and U54CA274371. A.M. and J.F.F. are supported by U01CA200468 and R01CA299972. E.K, M.K, and J.F.F. are supported by Pancreatic Cancer North America (PCNA). L.P. is supported by Translational Genomics and Precision Medicine in Cancer Training Program (T32 CA217789) and TRIUMPH Fellowship in the CPRIT Training Program (RP210028). E.I., J.Z., C.M.S., S.H., and J.F.F. are supported by NCI grant U01CA239522.

## Conflict-of-Interest Disclosure

**Dr. Maitra is listed as an inventor on a patent licensed to Exact Sciences (An Abbott labs company).**

---

**Supplemental Table S2.** Transcripts comprising the composite Matrix Stiffness Signature.

| Encoding Gene | Composite Matrix Stiffness Signature |
| --- | --- |
| COL1A1 |  |
| COL1A2 |  |
| FN1 |  |
| POSTN |  |
| LOX |  |
| PLOD2 |  |
| ACTA2 |  |
| TAGLN |  |
| CTGF |  |
| CYR61 |  |
| FAP |  |
| SPAR |  |

**Supplemental Figure S1.**
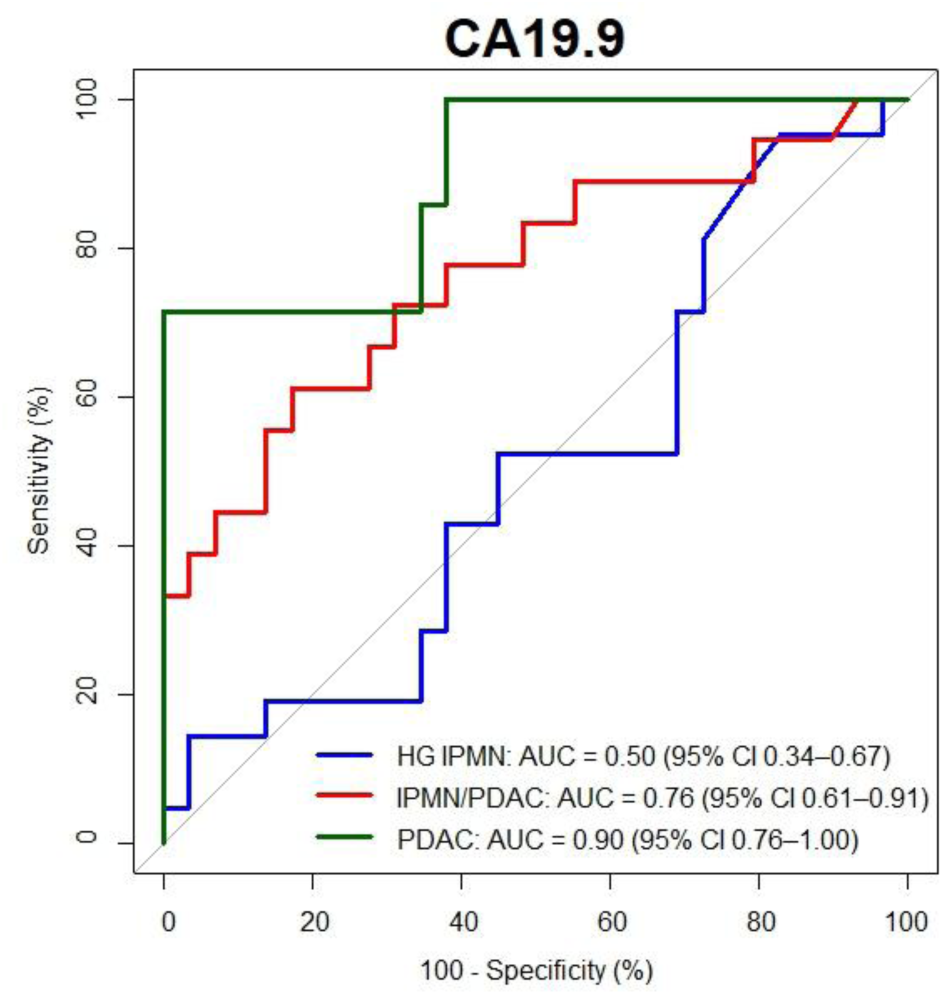
Predictive performance of CA19-9 for distinguishing HG IPMN, IPMN/PDAC, and PDAC without an IPMN from LG IPMN.

**Supplemental Figure S2.**
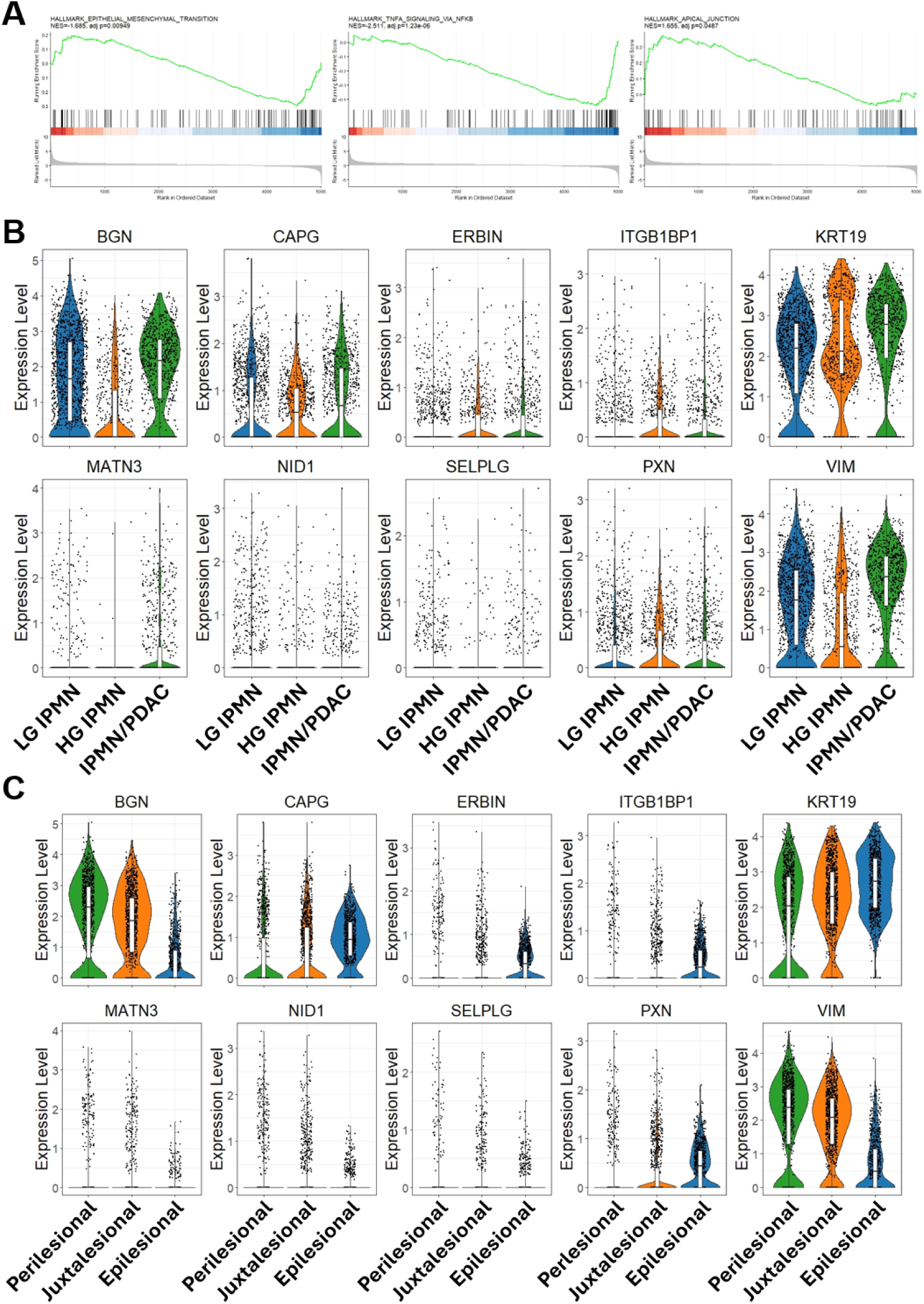
**A)** GSEA results for Hallmark EMT, TNFa signaling via NFkB, and Apical Junction based on DEGs (FDR-adjusted p-value <0.05) between HG IPMN and IPMN/PDAC tissues compared to LG IPMN tissues in the spatial transcriptomic dataset. **B)** mRNA expression levels for BGN, CAPG, ERBB2IP/ERBIN, ITGB1BP1, KRT19, MATN3, NID1, PSGL-1/SELPLG, PXN, and VIM in HG IPMN, IPMN/PDAC, and LG IPMN tissues. **C)** mRNA expression levels for BGN, CAPG, ERBB2IP/ERBIN, ITGB1BP1, KRT19, MATN3, NID1, PSGL-1/SELPLG, PXN, and VIM in epilesional, juxtalesional, and perilesional regions.

**Supplemental Figure S3.**
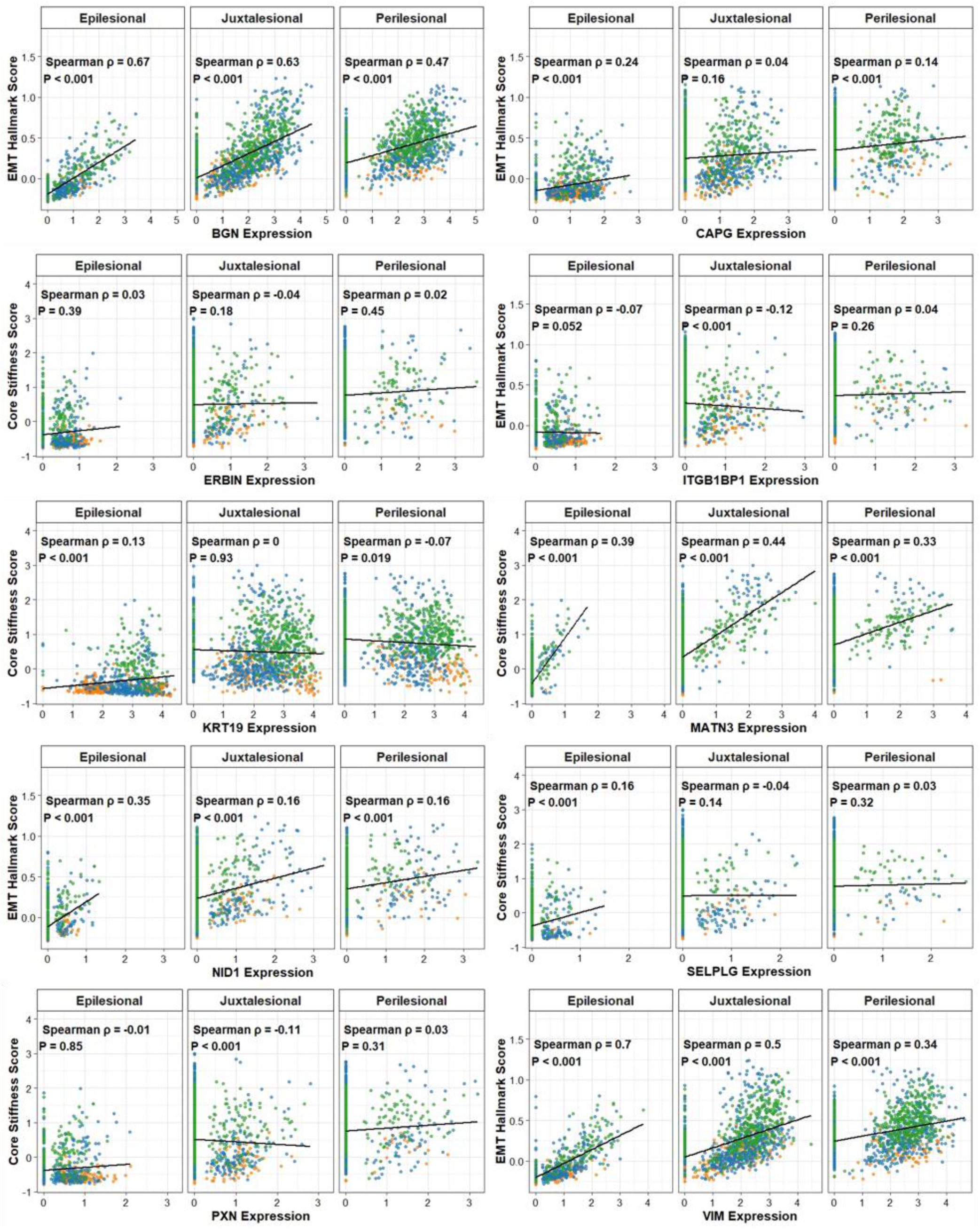
Scatter plots illustrating associations between BGN, CAPG, ITGB1BP1, NID1, and VIM with gene-based signature scores of matrix stiffness in epilesional, juxtalesional, and perilesional areas. Blue nodes- LG IPMN; Orange- HG IPMN; Green- IPMN/PDAC.

**Supplemental Figure S4.**
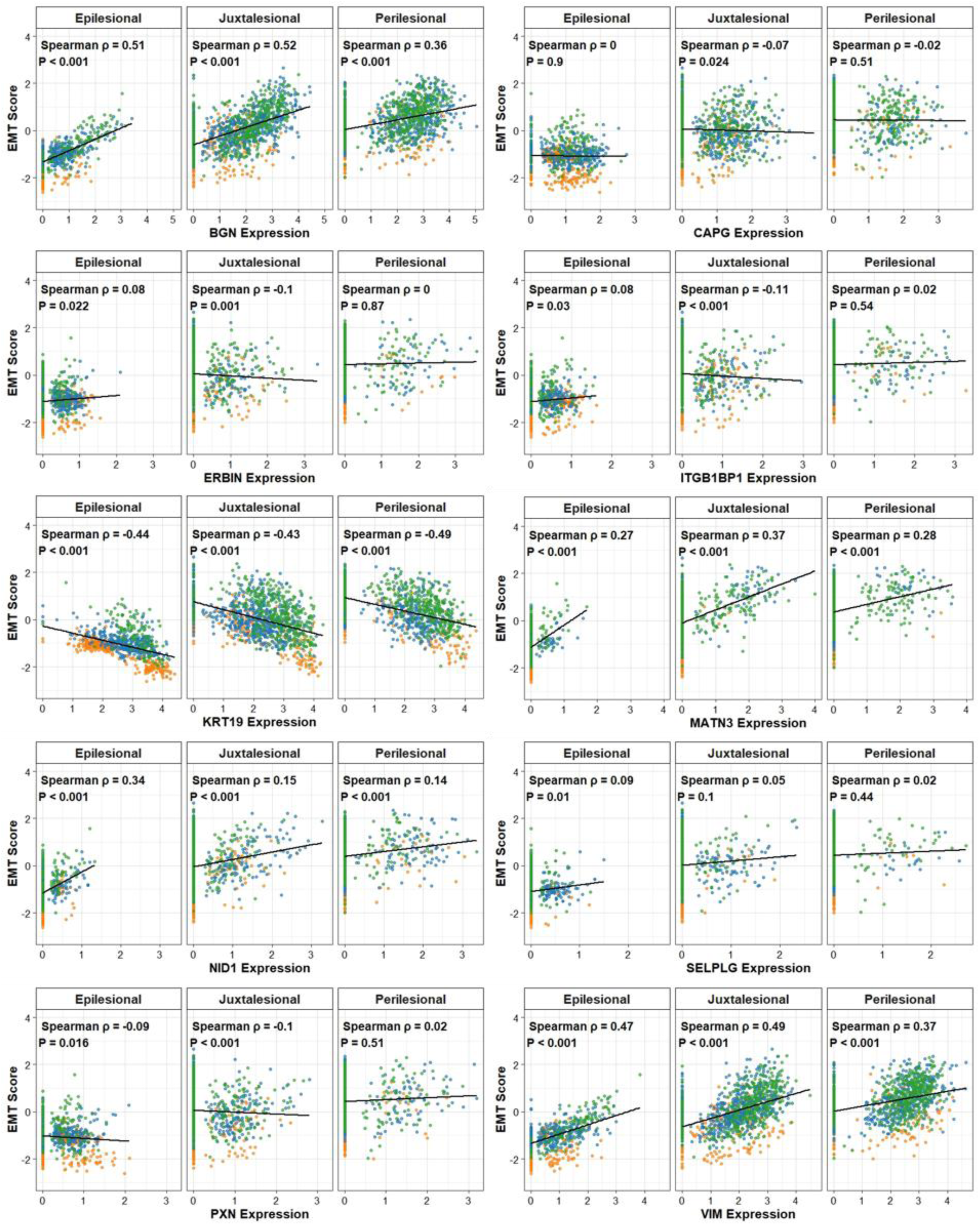
Scatter plots illustrating associations between BGN, CAPG, ITGB1BP1, NID1, and VIM with gene-based EMT signature scores in epilesional, juxtalesional, and perilesional areas. Blue nodes- LG IPMN; Orange- HG IPMN; Green- IPMN/PDAC.

**Figure S5.**
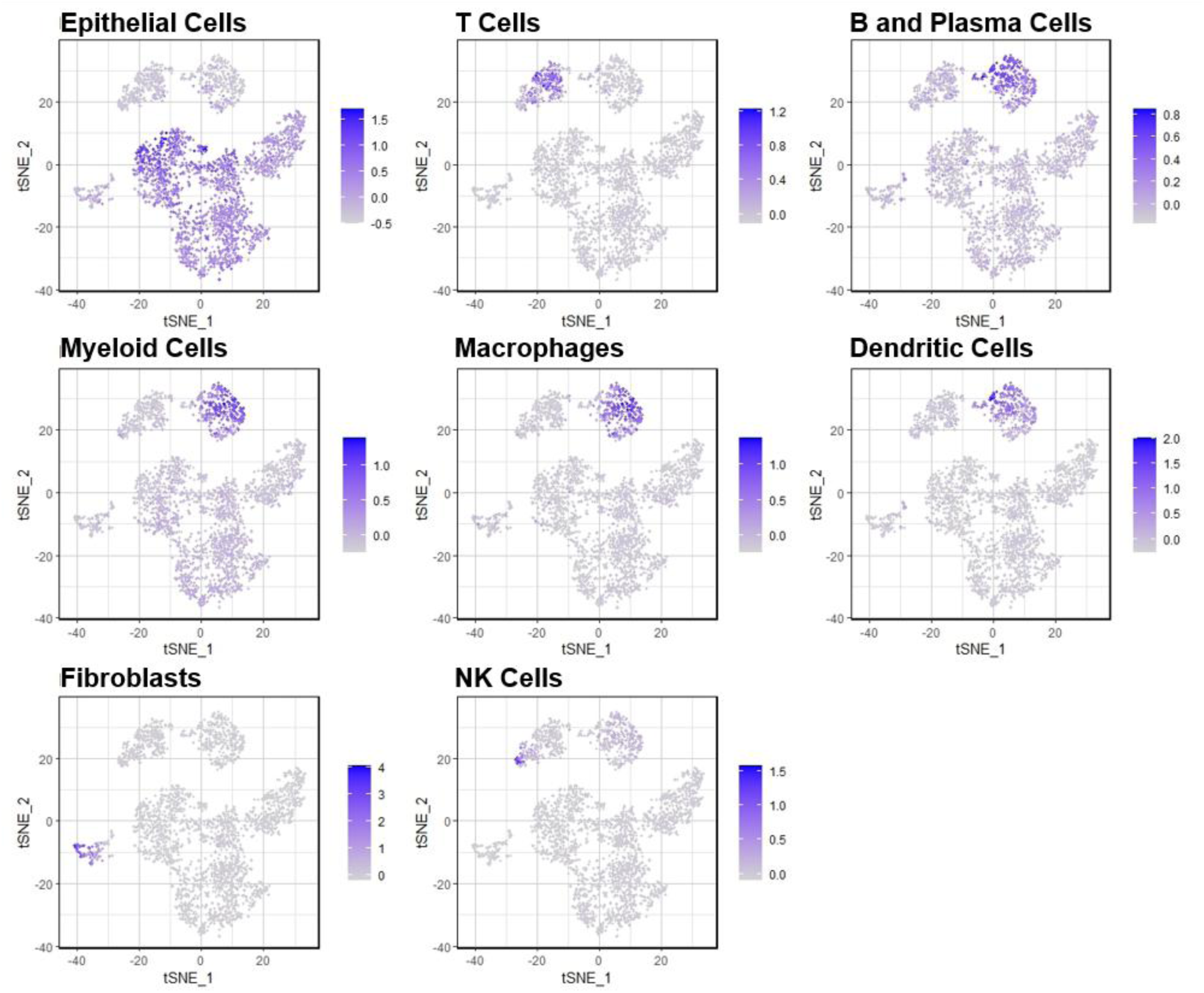
Representative tSNE plots for each of the 8 identified cell type populations in the IPMN single-cell transcriptomic dataset.

## Notes

### Author Declarations

Ethics committee/IRB of Indiana University School of Medicine gave ethical approval for this work. Ethics committee/IRB of The University of Texas MD Anderson Cancer Center gave ethical approval for this work.

